# Knowledge, attitude and practice of clinical audit among healthcare professionals in Gusau, Zamfara State, Nigeria

**DOI:** 10.64898/2025.12.18.25342282

**Authors:** Usman Hosea Ojoh, Dalhatu Nura, Wambutda Rotshak Moses, Nuhu Nitte

**Author notes:** Corresponding author: Usman Hosea Ojoh.

## Abstract

**Background:** Clinical audit is a vital quality improvement tool in healthcare that enables frontline workers to evaluate their care practices against established standards. This study assessed the knowledge, attitude, and practice of clinical audit among frontline healthcare workers in selected hospitals in Gusau, Zamfara State.

**Methods:** A descriptive cross-sectional study was conducted between February to September 2024 among 410 frontline healthcare workers across four health facilities in Gusau, Zamfara State, using a quantitative method of data collection. Data was analyzed using SPSS version 20, with a CI of 95% and p-value of ≤ 0.05 considered statistically significant.

**Results:** The mean age of respondents was 36.6 +/- 7.3 years, with 180 (44%) aged 31-40 years. The overall level of knowledge of clinical audit was adjudged to be poor among 224 (54.6%) of participants.

**Doctors:** demonstrated marginally higher good knowledge scores compared to other cadres. Facility type was significantly associated with knowledge level (p=0.001), while age, gender, specialty, and years of experience showed no significant association. Respondents generally had positive attitudes toward clinical audit, but actual participation in audit practices was low across all cadres.

**Conclusions:** This study has demonstrated a suboptimal level of knowledge of clinical audit among frontline healthcare workers, with variation across facility types. While attitudes toward clinical audit were largely positive, practical engagement remains limited, highlighting the need for targeted training and institutional support to enhance clinical audit practice in similar settings.

## Introduction

Clinical audit is a fundamental component of clinical governance, designed to enhance patient care and outcomes by systematically reviewing care practices against established criteria and implementing changes where necessary [1]. This process is critical in bridging the gap between recommended clinical practices and care patients receive, a discrepancy that often extends beyond patient-specific factors [2,3]. Clinical audits provide an objective means for healthcare professionals to evaluate their performance, as self-assessment can be inherently biased and challenging without external oversight [4]. Clinical audits have demonstrated significant improvements in healthcare quality and patient outcomes. For instance, in the United Kingdom, national clinical audits have led to marked reductions in healthcare inequalities and improvements in the management of conditions such as stroke [5,6]. However, the implementation and utilization of clinical audits in resource-poor settings, such as Nigeria, remain suboptimal [7]. While clinical audits have shown promise in improving maternal care and outcomes in similar settings, their adoption in Nigeria has been hindered by organizational, financial, and motivational challenges [8,9]. The knowledge and attitude of clinical audit among healthcare workers in Nigerian healthcare setting remain is largely limited. A recent study in Nigerian Tertiary hospital reveals concerning gap in clinicians understanding and application of clinical audit process, however these findings are constrained by sole focus on doctors neglecting other vital healthcare workers who contribute to patient care quality [10].

Furthermore, healthcare professionals often hold mixed attitudes toward clinical audit, with some viewing them as valuable tool for improving healthcare quality, while others see audit as time consuming and bureaucratic. These perceptions are often shaped by workload, institutional support and prior audit experience [11,12] Clinical audits effectiveness in resource-poor settings has been demonstrated for example, studies in Uganda and other developing countries which showed that criteria-based audits can significantly improve the quality of obstetric care, even in environments with limited resources [8,9]. These findings suggest that with the right strategies and support, clinical audits can be a powerful tool for improving healthcare quality in Nigeria. Current evidence regarding healthcare professionals’ knowledge, attitudes, and practices toward clinical audits in Nigeria’s health system remains limited, particularly among frontline providers who play a pivotal role in implementation. This knowledge gap underscores the need for a study assessing Knowledge, attitude and practice related to clinical audits among healthcare workers in different health facilities in Gusau, Zamfara State. Understanding these factors is critical for improving audit adoption and enhancing quality of care in similar setting. Materials And Methods

## METHODOLOGY

### Study setting

This study was conducted in four hospitals: Federal Medical Centre Gusau, Ahmad Yariman Specialist Hospital Gusau, King Farhad Women and Children Hospital, and Dr Karima Primary Healthcare Centre in Gusau, Zamfara State. Gusau is the capital of Zamfara State, located North of the line drawn from Kebbi to Sokoto. It has an estimated population of 682,700 and an area of 3,364km [13].

### Study population

The study population consisted of doctors, nurses, pharmacists, physiotherapists, community health extension workers, and pharmacists in all clinical departments’ units of Federal Medical Centre Gusau, Ahmad Yarima Specialist Hospital Gusau, King Farhad Women and Children Hospital, and Dr Karima Primary Healthcare Centre in Gusau, Zamfara State.

### Study Design

This study employed a cross-sectional descriptive design, conducted between February to September 2024, to determine the knowledge, attitude, and practice of clinical audit among health care workers across four selected healthcare facilities in Gusau, Zamfara State: Federal Medical Centre Gusau, Yariman Bakura Specialist Hospital Gusau, king Farhad women and children hospital Gusau, and Dr. Karima Primary Health Care Centre, using a quantitative method of data collection.

### Sample size estimation

The sample size for this study was calculated using the appropriate sample size determination formula for a cross sectional study n=Z ^2^ (1-p)/d ^2^ [14] Where n is the minimum sample size, Z is the standard normal deviate at 95% confidence interval (1.96), q is the complementary probability (1-p), d is the precision of the study set at 0.05 and p is the assumed (50%) of respondents with good knowledge. This gave a sample size of 422 after addition of 5% to cater for non or incomplete responses.

### Criteria for inclusion in the study

All doctors, nurses, pharmacist, physiotherapist, pharmacist and community health extension workers full time employees of the hospital, present at the time of the study and had spent at least one year on the jobs who had given consent for participation were included in the study. One (1) year period was set as cut-off for inclusion

### Study Design and Sampling Technique

The target population comprised of frontline health care workers including doctors, nurses, pharmacists, physiotherapists, and community health extension workers (CHEWs). A non-probability sampling technique was utilized to recruit participants. Data was collected through an online self-administered questionnaire, which was distributed among eligible health care workers using convenience and snowball sampling methods. Initial participants were identified through departmental 2 of 12 contacts and professional networks. The survey link was disseminated via hospital WhatsApp groups and direct peer referrals to enhance coverage. Eligible respondents were health care workers currently practicing in any of the selected facilities. Participation was voluntary, and informed consent was obtained. To improve participation, periodic reminders were sent, and data collection was conducted over a period of six months. A total of 422 responses were received and included in the final analysis.

### Data Collection Instrument

A semi-structured self-administered questionnaire adapted from previous studies [10] was used to collect data from participants. The questionnaire consisted of four sections: socio-demographic characteristics, knowledge of clinical audit, attitudes towards clinical audit, and practice of clinical audit among healthcare workers. The data collection instrument was pretested using both online and a printed version of the questionnaire at Yariman Bakura Specialist Hospital, involving 10% of the calculated minimum sample size. This pretest helped identify any ambiguities in the questions, allowed for the estimation of administration time, and ensured the instrument was appropriate for the study’s objectives. Grading of response A total of 7 questions were used to assess the level of knowledge, with 2 marks was allotted for each correct response and 0 mark for incorrect responses, giving a total attainable score of 14 with score equal to or greater than 8 considered as good knowledge. The distribution of scores was then categorized using a frequency table

### Data Analysis

The data obtained were processed, cleaned using excel removing greater than 50% incompletely filled responses given a total of 410 responses and analyzed using SPSS version 20. The socio-demographic characteristics of the respondents, such as age group, sex, marital status, etc., were expressed in terms of frequency and percentage. For continuous variables, the mean ± standard deviation was used as summary indices for age. The outcome variable in the study was the level of knowledge of clinical audit, categorized into good and poor knowledge, and presented as frequency and percentage. Assessment of the association between socio-demographic characteristics and knowledge scores, chi-square tests were employed. Cross-tabulation was performed to examine the relationship between socio-demographic factors and knowledge scores. A p-value of less than 0.05 was considered statistically significant, with a 95% confidence interval used for all statistical tests.

## Results

Table 1: Majority of respondents were aged 31-40 years (44%), with a mean age of 36.58 ± 7.3. Females constituted 57% of the participants. The distribution across specialties was balanced, with 33% each for doctors and nurses, 5% for pharmacists, and 29% for other professionals. Most respondents had over five years of work experience (56%) and worked in secondary (42%) and tertiary (37%) healthcare facilities. Only 41% had additional postgraduate qualifications.

**TABLE 1:** Socio-demographic characteristics of respondents.

| Variable | Knowledge Score | | | $\chi^2$ | P-Value |
| --- | --- | --- | --- | --- | --- |
|  | Good | Poor | Total |  |  |
| Age group |  |  |  | 28.441 | 0.737 |
| 21–30 | 47(45) | 58(55) | 105(26) |  |  |
| 31–40 | 78(44) | 101(56) | 179(44) |  |  |
| 41–50 | 48(48) | 53(52) | 101(25) |  |  |
| 51–60 | 9(36) | 11(64) | 25(5) |  |  |
| Gender |  |  |  | 0.400 | 0.527 |
| Male | 83(47) | 93(53) | 176(43) |  |  |
| Female | 103(44) | 131(55) | 234(57) |  |  |
| Specialty |  |  |  | 3.329 | 0.344 |
| Doctor | 68(50) | 67(50) | 135(33) |  |  |
| Nurse | 58(42) | 79(58) | 137(33) |  |  |
| Pharmacist | 6(32) | 13(68) | 19(5) |  |  |
| Others | 54(45) | 65(55) | 119(29) |  |  |
| Facility |  |  |  | 20.130 | 0.001 |
| Primary | 26(30) | 61(70) | 87(21) |  |  |
| Secondary | 72(42) | 100(58) | 172(42) |  |  |
| Tertiary | 87(58) | 63(42) | 150(37) |  |  |
| Additional academic qualification |  |  |  | 8.367 | 0.301 |
| Yes | 92(49) | 94(51) | 186(45) |  |  |
| No | 94(42) | 130(58) | 224(55) |  |  |
| Duration of practice (Years) |  |  |  | 25.611 | 0.817 |
| 1–5 | 70(39) | 110(61) | 180(44) |  |  |
| >5 | 110(48) | 120(52) | 230(56) |  |  |

Figure 1. : The knowledge scores regarding clinical audit among healthcare professionals, stratified into two categories: good and poor knowledge. This study reveal that a majority of respondents 54.6% exhibited poor knowledge, while 45.4% demonstrated good knowledge. Despite the overall poor knowledge score, doctors showed a marginally higher performance, with 50% achieving a good knowledge score.

**Figure 1.**
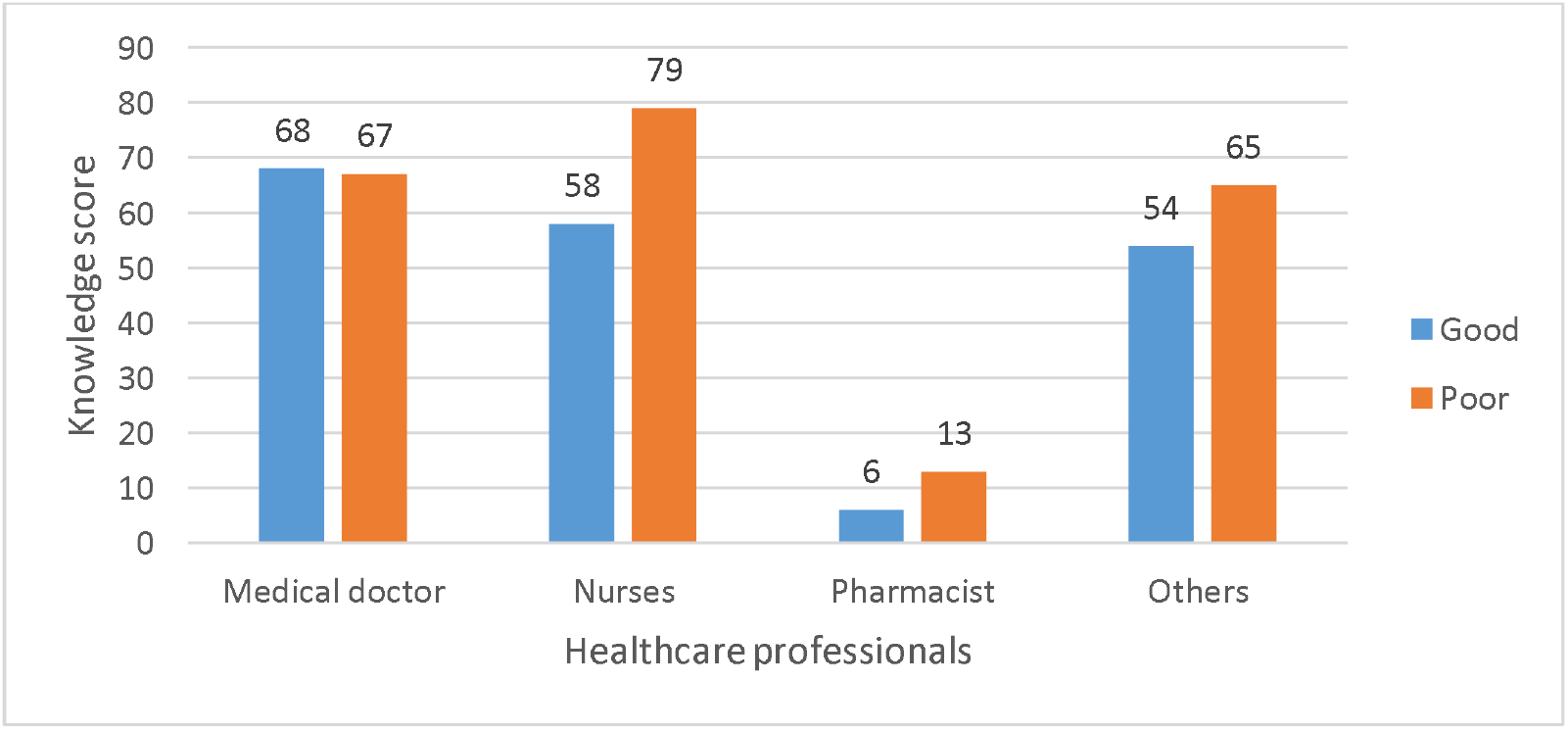
knowledge score on clinical audit among healthcare professionals

Table 2: The relationship between sociodemographic characteristics and clinical audit knowledge score of clinical audit practice amongst healthcare professionals showed a positive association with facility type (χ^2^ = 20.130, p = 0.001) with majority of participation 172(42) in secondary healthcare facility. No statistically significant associations between knowledge scores and age (χ^2^ = 28.441, p = 0.737), gender (χ^2^ = 0.400, p = 0.527), or professional specialty (χ^2^ = 3.329, p = 0.344) as well as other correlated variables.

**Table 2:**
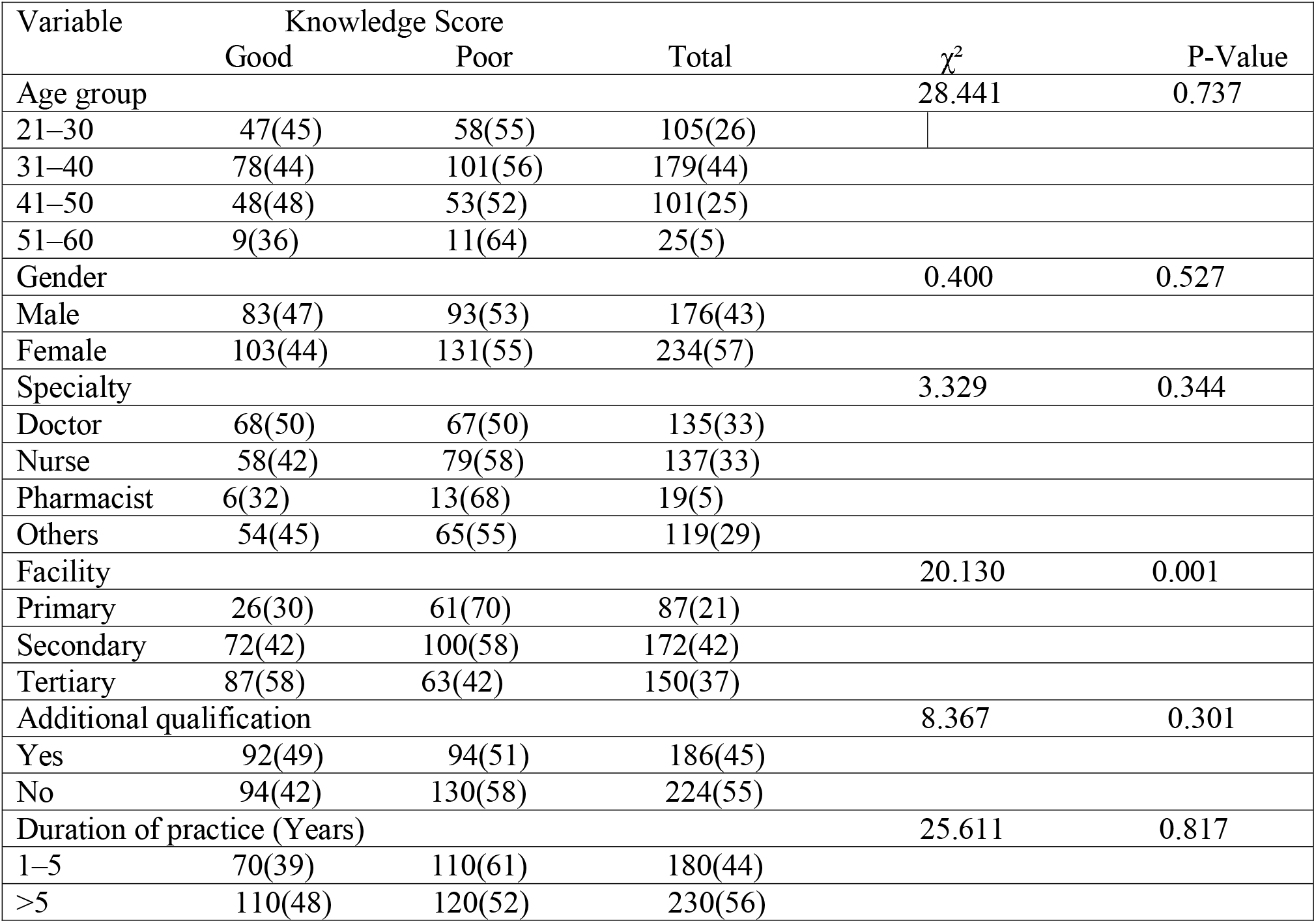
Association between Sociodemographic Characteristics and Knowledge Score of Clinical Audit among Healthcare Professionals.

| Variable | Knowledge Score | | Total | $\chi^2$ | P-Value |
| --- | --- | --- | --- | --- | --- |
|  | Good | Poor |  |  |  |
| Age group |  |  |  | 28.441 | 0.737 |
| 21–30 | 47(45) | 58(55) | 105(26) |  |  |
| 31–40 | 78(44) | 101(56) | 179(44) |  |  |
| 41–50 | 48(48) | 53(52) | 101(25) |  |  |
| 51–60 | 9(36) | 11(64) | 25(5) |  |  |
| Gender |  |  |  | 0.400 | 0.527 |
| Male | 83(47) | 93(53) | 176(43) |  |  |
| Female | 103(44) | 131(55) | 234(57) |  |  |
| Specialty |  |  |  | 3.329 | 0.344 |
| Doctor | 68(50) | 67(50) | 135(33) |  |  |
| Nurse | 58(42) | 79(58) | 137(33) |  |  |
| Pharmacist | 6(32) | 13(68) | 19(5) |  |  |
| Others | 54(45) | 65(55) | 119(29) |  |  |
| Facility |  |  |  | 20.130 | 0.001 |
| Primary | 26(30) | 61(70) | 87(21) |  |  |
| Secondary | 72(42) | 100(58) | 172(42) |  |  |
| Tertiary | 87(58) | 63(42) | 150(37) |  |  |
| Additional qualification |  |  |  | 8.367 | 0.301 |
| Yes | 92(49) | 94(51) | 186(45) |  |  |
| No | 94(42) | 130(58) | 224(55) |  |  |
| Duration of practice (Years) |  |  |  | 25.611 | 0.817 |
| 1–5 | 70(39) | 110(61) | 180(44) |  |  |
| >5 | 110(48) | 120(52) | 230(56) |  |  |

Table 3: Respondents generally held positive attitudes toward clinical audit, with the highest agreement on improving patient care (42.4% agreed), improving patient satisfaction (47% agreed), and using resources (40% agreed). However, concerns were noted regarding Diminished clinical ownership (32.9% agreed), Restricted clinical freedom (35.1% agreed), and Obstacles to individualized care (32.7% agreed). Institutional factors influencing clinical audit were mainly Clear policies on clinical audit (42.7% agreed), Availability of a central coordinating structure (40.2% agreed) and Time constraints for conducting audits (40.7% agreed). Only 39% of doctors, 29.7% of nurses, 26.2% of others, and 5.2% of pharmacists reported clinical audit activities in their departments.

**TABLE 3:**
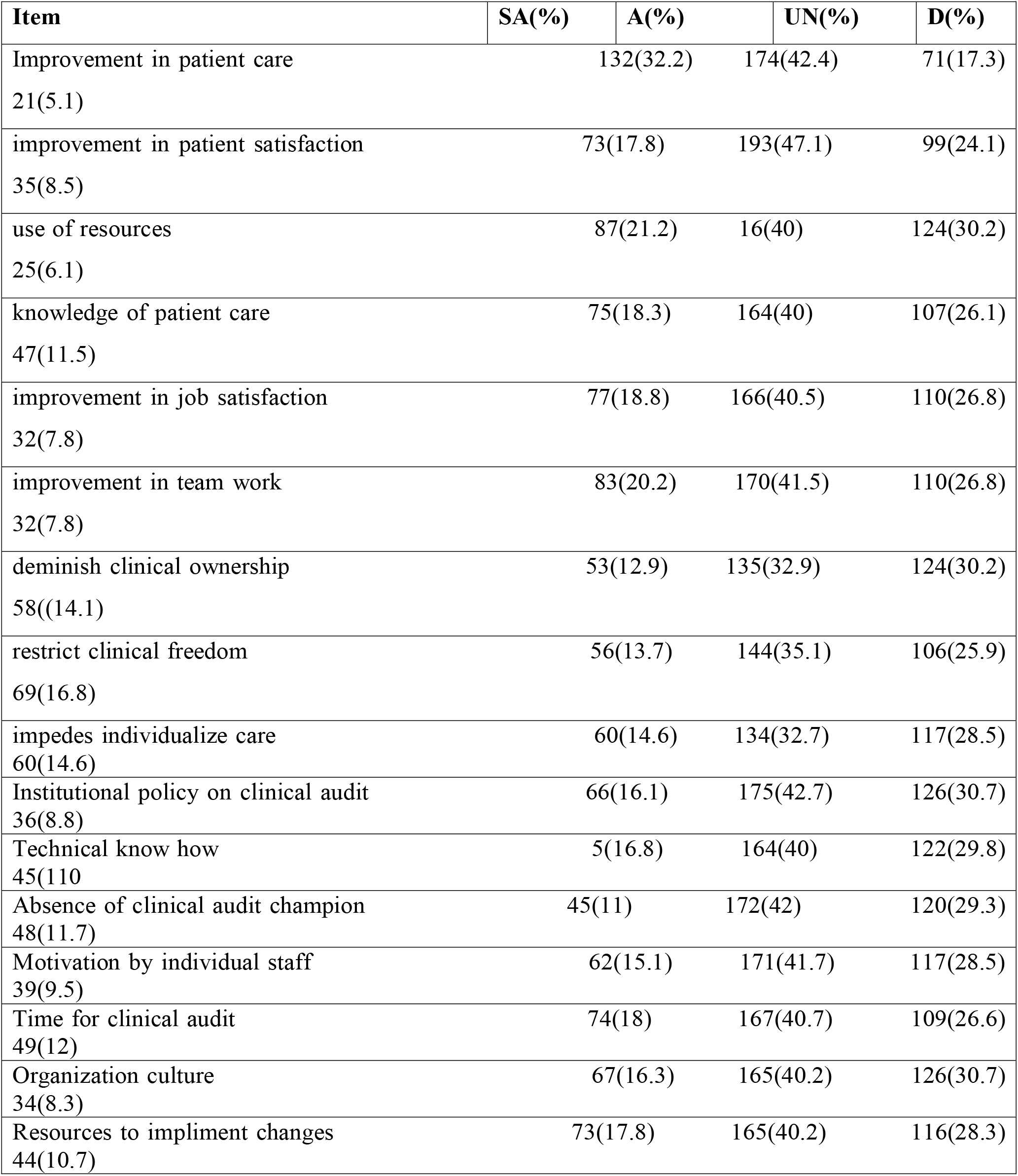
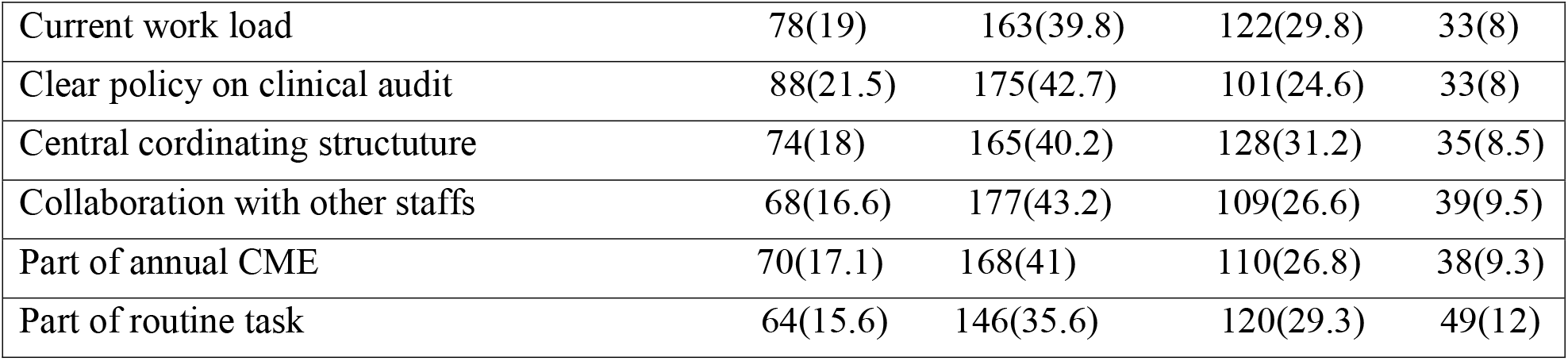
Attitude of healthcare professionals towards clinical audit.

Table 4: Among those trained, identifying audit needs (45% doctors, 27.5% nurses) and setting standards (57.1% doctors, 28.6% nurses) were the most covered aspects. Involvement in clinical audits was limited 39.1% of doctors, 29.3% of nurses, 28.8% of others, and 2.8% of pharmacists had participated in audits. Mortality audits (53.8% doctors, 27.5% nurses) and patient satisfaction audits (40% nurses, 38.2% others)

**TABLE 4:**
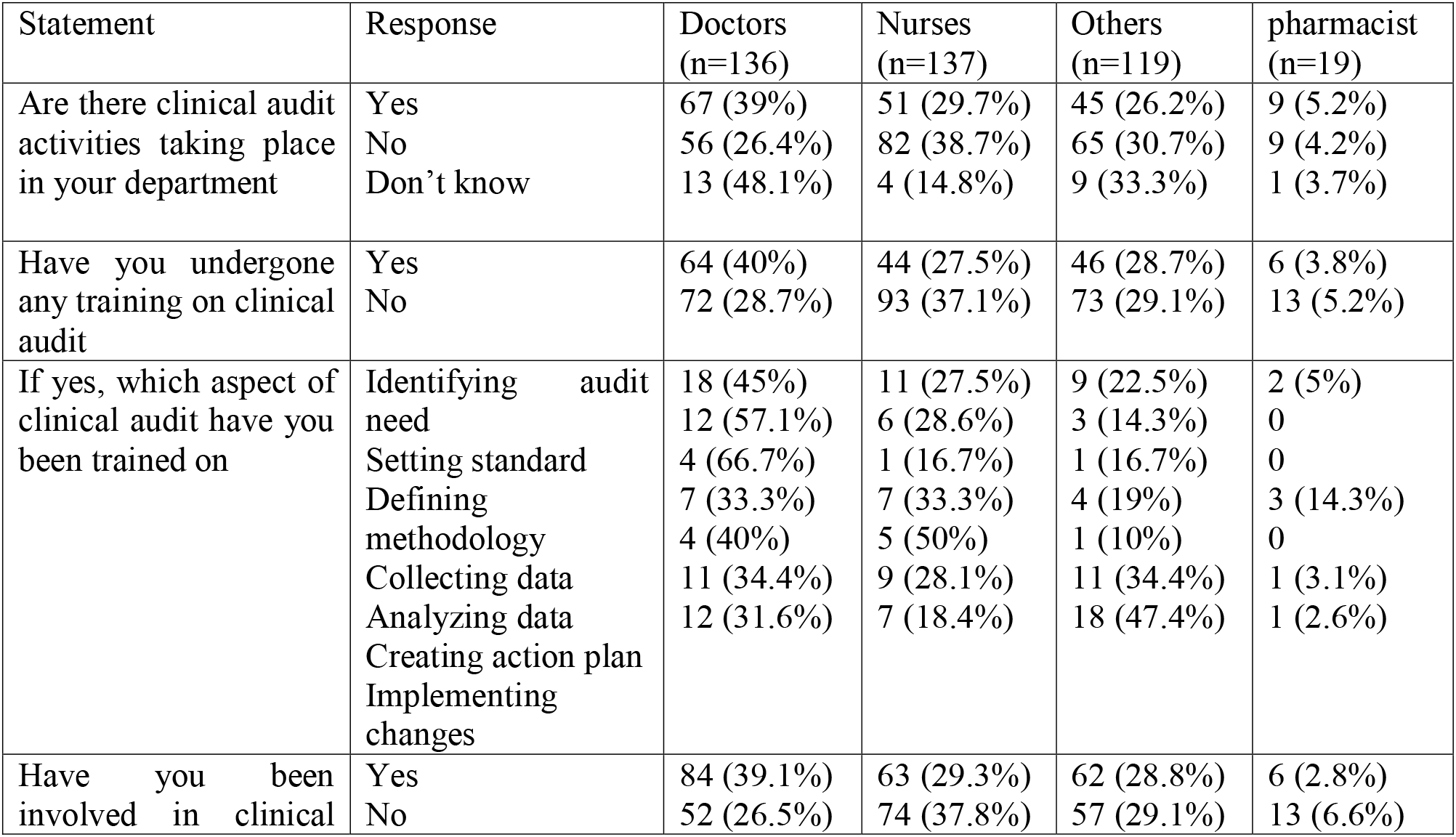
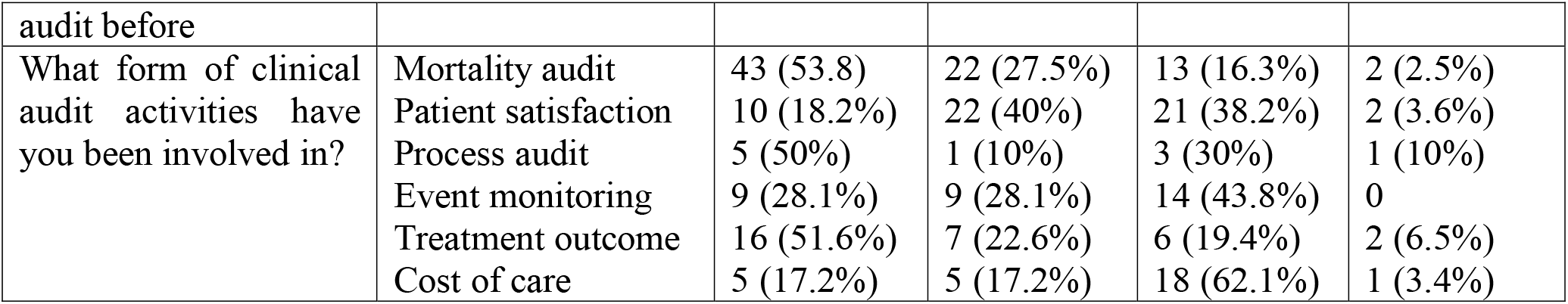
Practice of clinical audit among healthcare professionals.

Training on clinical audit was low, with 40% of doctors, 27.5% of nurses, 28.7% of others, and 3.8% of pharmacists having received any training.

## Discussion

Clinical audit serve as a vital tool for improving healthcare delivery by ensuring that clinical practices align with established standards [1]. Essentially, these study facilities lack an organized and working clinical audit system; therefore, it is imperative to assess healthcare workers’ knowledge, attitude, and practice of clinical audit as its utilization upon execution can be hinged on the knowledge base of this workforce. Understanding of clinical audit is fundamental to improving healthcare quality and patient outcomes. In this study, 54.6% of respondents demonstrated poor knowledge of clinical audit, while 45.5% showed good 7 of 12 knowledge. Notably, doctors had a marginally higher knowledge score of 50% compared to other frontline healthcare workers, though the overall knowledge level was suboptimal. This study is in synergy with a study in Kuwait [15] where doctors showed a good knowledge of clinical audit which can be attributed to training exposure to clinical audit, contrary with a study in Nigeria [10] where only 12.5% of doctors had a good knowledge of clinical audit. These two cited studies findings difference and limitations can be attributable to the fact that this study was conducted among medical doctors excluding other frontline healthcare workers and other healthcare facility type. A significant association [p-0.01,CI 95] as observed between knowledge level and facility type. This may be attributed to better access to resources, training, and institutional support in these facilities as they are designated to provide specialist care, this is similar to findings from studies in Uganda and Nigeria which showed the central role stake holders play in audit success [9,10]. Despite the knowledge gap, respondents generally had positive attitudes towards clinical audits. Most agreed that audits improve patient care, satisfaction, and efficient use of resources. These perceptions are consistent with several studies findings on the value of audits in clinical governance [3,7]. However, concerns about restricted clinical freedom and diminished autonomy were common and suggest the need for better communication on the developmental not punitive nature of audit [4]. In terms of practice, participation in audit activities was limited. Only a small proportion had received formal training, with doctors having higher level of involvement. This reflects challenges noted in previous research, where audit practice is often hindered by a lack of training and institutional support [9,10]. Institutional barriers such as unclear policies, time constraints, and absence of coordinating structures were they major constraints to effective audit practice. These findings are in line with literature that emphasizes the need for supportive environments to enable regular audit cycles. This study demonstrates positive attitude towards clinical audit among healthcare professionals, this provide a strong foundation for future interventions. Developing audit training programs, integrating audit topics into professional education, and establishing audit committees could help bridge the gap between knowledge and practice. The findings from this study provide useful insights for planning and implementing clinical audit strategies in comparable settings. To ensure effectiveness, interventions should be tailored to account for the specific roles and varying levels of experience among healthcare professionals.

## Limitations of study

This study utilized self-administered questionnaires, which may have introduced response bias, and employed a cross-sectional descriptive design that limits the ability to draw causal conclusions.

## Conclusions

This study highlights a notable gap in the knowledge and practical application of clinical audits among frontline healthcare workers in Gusau, Zamfara State. While a positive attitude toward clinical audits was observed across all professional cadres, over half of healthcare professionals demonstrated inadequate knowledge, and participation in audit activities was limited. Doctors exhibited marginally better knowledge, clinical audit knowledge level was significantly associated with the type of facility, suggesting institutional context plays a key role in exposure and training.

## Data Availability

Available with the principal author and Institution where ethical clearance was given and Available on Request

## Additional Information Disclosures

### Human subjects

Consent for treatment and open access publication was obtained or waived by all participants in this study. Research and ethics committee issued approval AYSBSH/SUB/205/VOL.1. Animal subjects: All authors have confirmed that this study did not involve animal subjects or tissue.

### Conflicts of interest

In compliance with the ICMJE uniform disclosure form, all authors declare the following:Payment/services info: All authors have declared that no financial support was received from any organization for the submitted work. Financial relationships: All authors have declared that they have no financial relationships at present or within the previous three years with any organizations that might have an interest in the submitted work. Other relationships: All authors have declared that there are no other relationships or activities that could appear to have influenced the submitted work.

## Notes

### Competing Interest Statement

The authors have declared no competing interest.

### Funding Statement

No funding was provided for this research

### Author Declarations

Informed consent for treatment and open access publication was obtained or waived by all participants in this study. Research and Ethics Committee issued approval in Yariman Bakura Specialist Hospital Gusau, Research and Ethics Committee AYSBSH/SUB/205/VOL.1

## References

1. Charles D. Shaw: Principles for Best Practice in Clinical Audit. International Journal for Quality in Health Care. 2003, 15:87–087. 10.1093/intqhc/15.1.87

2. McPherson K: Why do variations occur? In: Andersen TF, Mooney G, editors. The Challenges of Medical Practice Variations. London: Palgrave. 2002, 23–45. 10.1007/978-1-349-20781-7_2

3. Skull S: Embedding clinical audit into everyday practice: essential methodology for all clinicians. J Paediatr Child Health. 2020, 56:1533–1536. 10.1111/jpc.15068

4. Colthart I, Bagnall G, Evans A, et al.: The effectiveness of self-assessment on the identification of learner needs, learner activity, and impact on clinical practice. BEME Guide no. 10. Med Teach. 2008, 30:124-145.10.1080/01421590701881699

5. Hanskamp-Sebregts M, Zegers M, Boeijen W, et al.: Effects of auditing patient safety in hospital care: design of a mixed-method evaluation. BMC Health Serv Res. 2013, 13:226–10. 10.1186/1472-6963-13-226.

6. Leading the information revolution in cancer intelligence: why the National Lung Cancer Audit is the key to transforming lung cancer outcomes. (2014). Accessed: 22 May 2025:http://documents.roycastle.org/leading-the-information-revolution-in-cancer-intelligence.pdf.

7. Hussein J, Hirose A, Owolabi O, et al.: Maternal death and obstetric care audits in Nigeria: a systematic review of barriers and enabling factors in the provision of emergency care. Reprod Health. 2016, 13:47–10. 10.1186/s12978-016-0158-4

8. KI Hunyinbo, AO Fawole, OS Sotiloye, et al. (Evaluation of criteria-based clinical audit in improving quality of obstetric care in a developing country hospital). Accessed: Afr J Reprod Health: https://pubmed.ncbi.nlm.nih.gov/19435013/.

9. Weeks AD, Alia G, Ononge S, et al.: Introducing criteria-based audit into Uganda maternity units. BMJ. 2003, 327:1329. 10.1136/bmj.327.7427.1329

10. Alinnor EA, Ogaji DS: Physicians’ knowledge attitude and practice of clinical audit in a tertiary health facility in a developing country: a cross-sectional study. Pan Afr Med J. 2022, 43:22. 10.11604/pamj.2022.43.22.33800

11. McWilliams S, Schofield S: What influences postgraduate psychiatric trainees’ attitudes to clinical audit?.Irish Journal of Psychological Medicine. 2020, 37:106–110. 10.1017/ipm.2017.6

12. Lord J, Littlejohns P: Impact of hospital and community provider based clinical audit programmes: perceptions of doctors, nurses and other health professionals. Int J Qual Health Care. 1996, 8:527–535.

13. Gusau local Government area. (2025). Accessed: 18 May: https://zamfara.gov.ng/elementor-331/.

14. Ahmed SK: How to choose a sampling technique and determine sample size for research: a simplified guide for researchers. Oral Oncol Rep. 2024, 12:100662. 10.1016/j.oor.2024.100662

15. Al-Baho A, Serour M, Al-Weqayyn A, et al.: Clinical audits in a postgraduate general practice training program: an evaluation of 8 years’ experience. PLoS One. 2012, 7:43895. 10.1371/journal.pone.0043895

